# Multi-level modeling and multiple group analysis of disparities in continuity of care and viral suppression among Nigerian adolescents and youths living with HIV

**DOI:** 10.1101/2020.06.28.20141812

**Authors:** Okikiolu Abimbola Badejo, Christiana Noestlinger, Toyin Jolayemi, Juliet Adeola, Prosper Okonkwo, Sara Van Belle, Edwin Wouters, Marie Laga

## Abstract

**Introduction:** Substantial disparities in care outcomes exist between different sub-groups of adolescents and youths living with HIV(ALHIV). Understanding variation in individual and health-facility characteristics could be key to identifying targets for interventions to reduce these disparities. We modeled variation in ALHIV retention in care and viral suppression, and quantified the extent to which individual and facility characteristics account for observed variations.

**Methods:** We included 1,177 young adolescents (10-14 years), 3,206 older adolescents (15-19 years) and 9,151 young adults (20-24 years) who were initiated on antiretroviral therapy (ART) between January 2015 and December 2017 across 124 healthcare facilities in Nigeria. For each age-group, we used multilevel modeling to partition observed variation of main outcomes(retention in care and viral suppression at 12 months post ART initiation) by individual (level one) and health facility (level two) characteristics. We used multiple group analysis to compare the effects of individual and facility characteristics across age-groups.

**Results:** Facility characteristics explained most of the observed variance in retention in care in all the age-groups, with smaller contributions from individual-level characteristics (14-22.22% vs 0 - 3.84%). For viral suppression, facility characteristics accounted for a higher proportion of variance in young adolescents (15.79%), but not in older adolescents (0%) and young adults(3.45%). Males were more likely to not be retained in care(aOR=1.28; p<0.001 young adults) and less likely to achieve viral suppression (aOR=0.69; p<0.05 older adolescent). Increasing facility-level viral load testing reduced the likelihood of non-retention in care, while baseline regimen TDF/3TC/EFV or NVP increased the likelihood of viral suppression.

**Conclusions:** Differences in characteristics of healthcare facilities accounted for observed disparities in retention in care and, to a lesser extent, disparities in viral suppression. An optimal combination of individual and health-services approaches is, therefore, necessary to reduce disparities in the health and wellbeing of ALHIV.

**Key Messages:** *What is already known?:*

- Adolescents and youths living with HIV have worse care and treatment outcomes compared to other groups
- Significant disparity in care and treatment outcomes exist between different adolescent and youth subgroups

*What are the new findings?:*

- Differences in characteristics of healthcare facilities delivering services are associated with disparities in outcomes within and across adolescent and youth age-groups.

*What do the new findings imply?:*

- An optimal combination of individual and health-services approaches is necessary to reduce disparities in the health and wellbeing of adolescent and youths living with HIV.

## 1. Introduction

Adolescents and youths are an important group in global efforts to eliminate HIV. Compared to adults, adolescents and youths engage less with health care services and have lower viral suppression rates (1–5), resulting in increases in AIDS-related deaths despite a global reduction of AIDS-mortality among other age-groups (6–8). Under current circumstances, the global optimism to eliminate HIV by 2030 does not appear to hold for adolescents and youths (9,10).

Sub-optimal outcomes among adolescents and youths living with HIV (ALHIV) has been ascribed to unique individual vulnerabilities due to developmental factors associated with transitioning to adulthood (11–13). However, recent research suggests that such poor outcomes also demonstrate a failure of clinical care services to address the unique needs of ALHIV as they transition to adulthood. Research in this area seeks to identify and modify aspects of health services to be more adolescent and youth-friendly to improve care and treatment outcomes. Although some of these efforts highlight some promising strategies results have been mixed (14–24). More important, much of the research does not sufficiently disentangle the multi-layered influences of individual- and health services factors. This is important because delineating the relative contributions of individual and higher-level factors can inform the design of specific individual- or higher-level interventions across often different and heterogeneous ALHIV groups and communities.

In this multilevel study, we, therefore, examined the interplay of factors influencing ALHIV outcomes at the individual- and health-services levels as they relate to retention in care and viral suppression among a large sample of ALHIV in Nigeria. Nigeria accounts for a significant proportion of ALHIV burden in sub-Saharan Africa, and one of the few countries where mortality among ALHIV continues to increase (25). The national HIV program began in 2002 in only 25 public (government-owned) tertiary-level healthcare facilities. Gradual decentralization since then has resulted in an increased mix in types and levels of health care facilities delivering HIV services (26). While this has significantly increased access to HIV services, it has also brought about heterogeneity in terms of how national HIV guidelines are interpreted and implemented. The extent to which these differences in service delivery channels have shaped HIV-related outcomes have received little attention partly because of little information sharing between the public and private health sectors. Our study will characterize how this heterogeneity operates alongside individual-level characteristics to affect retention in care and viral suppression within and between ALHIV age-groups, thereby enabling the development of more context-appropriate individual or service-delivery interventions. We hypothesize that differences in characteristics of healthcare facilities delivering services to ALHIV will be associated with differences in these outcomes within and across ALHIV age-groups.

## 2. Methods

### Study setting and study population

Nigeria is administratively divided into 36 states and within these states, comprehensive HIV services are freely provided by the Government of Nigeria with funding support from the US President’s Emergency Plan for AIDS Relief (PEPFAR). Our study covered two of the 36 states (Benue and Plateau).

We retrospectively evaluated routine data for ALHIV enrolled in HIV care and treatment between 1 January 2015 to 31 December 2017 within a network of healthcare facilities supported by the APIN Public Health Initiatives Ltd/Gte treatment network (APIN). APIN is a PEPFAR implementing agency that provides technical and programmatic support to 285 health-care facilities to deliver comprehensive HIV care and treatment, spread across 95 local government areas (LGAs) in eight states of Nigeria (the country has a total of 774 LGAs distributed across 36 states; LGAs may be the equivalent of districts in other settings). The cohort for this study was enrolled in HIV care across 124 secondary and tertiary level healthcare facilities in two of the program’s eight states (Benue and Plateau states). Secondary facilities provide HIV services within a generalist, non-specialized clinic setting, while tertiary facilities provide services in specialist hospital settings. Healthcare facilities were included if they had provided comprehensive HIV prevention, care, and treatment services for at least one year at the time of the study.

### Enrollment into care and follow-up

All ALHIV in our study received free comprehensive HIV care according to Nigeria’s national guidelines for HIV prevention, care, and treatment, which is regularly updated according to the WHO recommendations. These include antiretroviral treatment (ART) for children, adolescents, and adults, the prevention of mother-to-child-transmission services (PMTCT); laboratory services including viral load, CD4 count, routine safety monitoring of laboratory test results; and pharmacy services.

### Outcome variables

#### Retention in care

To assess this outcome, we adapted indicators 1.2 and 1.3 of the Global AIDS Monitoring tool (GAM) (27). GAM indicator 1.2 describes the proportion of people living with HIV who are on antiretroviral therapy including people reinitiating antiretroviral therapy after previously having stopped treatment or being classified as lost to follow-up. We adapted this indicator to define retention according to the pattern of care-engagement at three-time points (30 days, 90 days, and 365 days) within the first 12 months after initiating ART. ALHIV were considered retained in care if they established care contact at all the three time points as evidenced by documented clinic visit and/or drug pick up. ALHIV were considered to have interrupted care if they missed care contact on one or two of the three time-points, and classified as lost to follow up (LTF) if there was no care contact at all three time-points after initiating ART.

#### Viral suppression

GAM indicator 1.3 assesses the percentage and number of adults and children living with HIV who have suppressed viral load and tracks progress towards the third 90 of UNAIDS 90-90-90 targets (28). For the numerator, the GAM recommends reporting the estimated number of people who have suppressed viral load if viral load testing coverage (i.e., the number of people routinely tested among all people on treatment) is ≥50%; or reporting only the number of routine viral load tests if viral load testing coverage is less than 50%. As the viral load coverage in our study population exceeded 50%, we adapted both the numerator (viral suppression rates) as our second outcome and the denominator (viral load testing coverage) as one of the explanatory variables at the service-delivery level (described below). Consistent with the GAM indicator 1.3 we defined viral suppression as HIV-RNA viral load < 1000 copies/ml. Although national policy recommends a viral load test at 12 months, we used viral load tests conducted within a 12-18 month window after ART initiation to allow some time for viral load testing among tracked ALHIV who returned to care following care interruption or LTF

### Exposure variables

#### Individual-level data measures

We included demographic and clinical information routinely collected at baseline during enrollment into HIV care. Demographic information collected included age at enrolment, sex, marital status, pregnancy status, level of education. Clinical characteristics included CD4 cells/ml count at enrollment, TB status, WHO clinical stage, ART regimen received. Information about the follow-up drug or clinic appointments visits were also available.

#### Facility-level measures

We used variables at the health care facility level to assess characteristics at the service delivery level. Such information included type/level of services offered (secondary or tertiary level care), the facility size (i.e. total number of ALHIV enrolled in care categorized as < 200, 200 - 500, > 500), viral load test coverage (proportion of enrolled ALHIV who have had at least one viral load test at 12 months or more after initiating ART) and whether the health facility was a designated adolescent/youth friendly center. Adolescents and youth centers within the APIN program refer to any healthcare facility implementing dedicated services to adolescents and youths in addition to routine HIV service provision. Such services are focused on modifying certain structures of care to be adolescent-friendly and may include re-organizing or changing the location of the clinic, dedicated clinic days, availability of peer support groups, adolescent–provider communication modalities, appointment availability and scheduling, and trained providers for adolescents and youth.

### Data management

We retrieved de-identified clinical data from the APIN/PEPFAR electronic database, which routinely captures patient demographic, clinical, and laboratory data. APIN supports health facility information systems with relational database management that is opensource (OpenMRS; www.openmrs.com).

### Statistical analysis

We first conducted descriptive analyses of baseline individual and facility-level characteristics using proportions, frequencies, means, and median. ALHIV in the cohort were categorized into three age-groups (10-14 years young adolescents, 15-19 years older adolescents, 20-24 years young adults) and the baseline characteristics were compared for the three age-groups using chi-square statistics. Next, we conducted a bivariate analysis of age-group comparisons of patterns of retention in care and viral measures.

To develop multi-level models to examine the association between individual- and facility-level factors and the two outcomes of interest (retention in care and viral suppression), we adopted a binary outcome logistic multilevel modeling approach, partitioning each outcome’s variance by its individual-(level one) and facility-level (level two) components concurrently to account for the nested structure of our data. In this approach, we allow the model intercepts to vary at random. We conducted the modeling analysis in three steps. In the first step (Model 1, null model) we did not include any explanatory variable, allowing an estimate of the total variation in outcomes by health facilities. In the second step (Model 2), we included only individual-level factors. This step allowed us to test whether or how much of the total random variation observed in the first stage could be explained by our individual-level data. In the third step (Model 3) we added facility-level factors to Model 2. This allowed us to test whether or how much of the random variation could be explained by the facility-level characteristics. For each outcome, we repeated these model steps for each of our three ALHIV age-group.

**Measures of association (fixed effects)** in our models are presented as odds ratios (ORs) and their p-values.

**Measures of variation (random effects)** in the models are random slope variance, intra-cluster correlation (ICC), and explained variation. Random slope variance indicates whether a contextual phenomenon differs in magnitude for different groups and whether the facility level modifies associations between individual-level exposures.

To check whether the effect of explanatory variables differed across ALHIV age-groups, we conducted a multiple group analysis. We used *mysuest* program to combine models from the three ALHIV age-groups and then used *mitesttransform* command to test for equality of each variable coefficients across the three age-groups using a 5% significance threshold.

To minimize potential bias due to missing data, we performed multiple imputations with chained equations after data exploration to verify that data were missing at random. A total of 163 data imputations were done as determined by the STATA module “how_many_imputations” (29) All statistical analyses were conducted using Stata version 16.1 (Stata Corporation, College Station, Texas, USA).

### Ethics

As part of the APIN protocol written informed consent and, where necessary, assent for service provision and participation (or data use) for future evaluations were obtained at the point of enrollment into HIV care and treatment. Adolescents or minors less than 15 years at enrolment provide another written and signed informed consent as soon as they become 15 years or older when transferred/transitioned to the adult ART program. Ethical approval for this study was obtained from the IRBs of APIN and the Institute of Tropical Medicine, Antwerp.

## 3. Results

### Baseline characteristics

Our study included a total of 13,527 AYLHIV who were initiated on ART between 1 January 2015 and 31 December 2017. Table 1 below presents the baseline characteristics.

**Table 1:** Baseline Characteristics of ALHIV by age-group

| Characteristics | Total | Young Adolescents<br>(10-14years) | Older Adolescents<br>(15-19years) | Young Adults<br>(20-24 years) | X p-value |
| --- | --- | --- | --- | --- | --- |
| AYLHIV (N, %) | 13,527(100%) | 1,177 (8.7) | 3,206 (23.7) | 9,151 (67.6) |  |
| Female, n (%) | 11,380 (84.1) | 656 (56.1) | 2,658 (82.9) | 8,066 (88.1) |  |
| **Age at enrolment**<br>**(Median, IQR)** | 21 (19-23) | 12 (10-13) | 18 (17-19) | 22(21-23) |  |
| Female | 21 (19-23) | 10 (12-13) | 18(17-19) | 22 (21-23) |  |
| Male | 20 (15-22) | 10(12-13) | 17 (16-18) | 22(21-24) |  |
| **Marital Status, n (%)** |  |  |  |  | 0.000 |
| Single | 1,481 (33.4) | 169 (82.4) | 492 (46.6) | 820 (25.9) |  |
| Married | 2,365 (53.4) | 18 (8.8) | 453 (42.9) | 1,894 (59.8) |  |
| Others | 581 (13.1) | 18 (8.8) | 111 (10.5) | 453 (14.3) |  |
| Missing | 9,100 | 965 | 2,150 | 5,985 |  |
| **Pregnancy status, n (% of females)** |  |  |  |  | 0.06 |
| Pregnant | 121 (1) | 1 (0.2) | 32 (1.2) | 88 (1.1) |  |
| Non-pregnant | 8,971 (99) | 130 (99.8) | 2,031 (98.8) | 6810 (98.9) |  |
| Missing | 2,288 | 525 | 595 | 1,168 |  |
| **Education, n (%)** |  |  |  |  | 0.00 |
| None/Informal | 1072 (34.6) | 45 (31) | 257 (33.7) | 770 (35.1) |  |
| Primary | 610 (19.7) | 60 (41.4) | 171 (22.4) | 379 (17.3) |  |
| Secondary and higher | 1418 (45.7) | 40 (27.6) | 334 (43.8) | 1044 (47.6) |  |
| Missing | 10,427 | 1025 | 2444 | 6958 |  |
| **Level of care, n(%)** |  |  |  |  |  |
| Secondary | 12658 (93.6) | 1041 (89) | 2940 (91.7) | 8677 (94.8) | 0.00 |
| Tertiary | 869 (6.4) | 129 (11) | 266 (8.3) | 474 (5.2) |  |
| **Year of ART initiation, n (%)** |  |  |  |  | 0.00 |
| 2015 | 4509 (33.3) | 343 (29.3) | 1037 (32.3) | 3129 (34.2) |  |
| 2016 | 4997 (36.9) | 430 (36.8) | 1233 (38.5) | 3334 (36.4) |  |
| 2017 | 4021 (29.7) | 397 (33.9) | 936 (29.2) | 2688 (29.4) |  |
| **Baseline CD4 cells/ml (Median, IQR)** | 345 (198-516) | 399 (185-634) | 348 (201-515) | 337 (198-508) |  |
| **Baseline immunodeficiency, n(%)** |  |  |  |  | 0.00 |
| None (CD4≥500cells/ml) | 1432 (26.9) | 157 (34.9) | 346 (27) | 929 (25.8) |  |
| Mild (CD4≥350-499cells/ml) | 1190 (22.3) | 100 (22.3) | 287 (22.4) | 803 (22.3) |  |
| Advanced (CD4≥200-349cells/ml) | 1364 (25.6) | 76 (16.9) | 332 (25.9) | 956 (26.6) |  |
| Severe (CD4 < 200cells/ml) | 1342 (25.2) | 116 (25.8) | 316 (24.7) | 910 (25.3) |  |
| Missing | 8199 | 721 | 1925 | 5553 |  |
| **TB Status at baseline, n (%)** |  |  |  |  | 0.27 |
| No signs or symptoms of TB | 7442 (92.7) | 589 (92.9) | 1794 (92.6) | 5059 (92.7) |  |
| Presumptive or confirmed TB | 443 (5.5) | 32 (5) | 100 (5.2) | 311 (5.7) |  |
| On IPT | 143 (1.8) | 13 (2.1) | 44 (2.3) | 86 (1.6) |  |
| Missing | 5499 | 536 | 1268 | 3695 |  |
| **WHO Stage, n(%)** |  |  |  |  | 0.00 |
| 1 | 6475 (77.7) | 493 (74) | 1584 (79.3) | 4398 (77.6) |  |
| 2 | 1272 (15.3) | 105 (15.8) | 274 (13.7) | 893 (15.8) |  |
| 3 | 517 (6.2) | 61 (9.2) | 119 (6) | 337 (5.9) |  |
| 4 | 67 (0.8) | 7 (1) | 21 (1) | 39 (0.7) |  |
| Missing | 5196 | 504 | 1208 | 3484 |  |
| **Baseline ART Regimen, n(%)** |  |  |  |  | 0.00 |
| ABC or AZT/3TC/EFV or NVP | 2048 (15.1) | 740 (63.2) | 466 (14.5) | 842 (9.2) |  |
| ABC or AZT/3TC/ATVr or LPVr | 38 (0.3) | 9 (0.8) | 16 (0.5) | 13 (0.1) |  |
| TDF/3TC/EFV or NVP | 11349 (83.9) | 409 (35) | 2691( 83.9) | 8249 (90.1) |  |
| TDF/3TC/ATVr or LPVr | 32 (0.2) | 2 (0.2) | 18 (0.6) | 12 (0.1) |  |
| Others | 60 (0.4) | 10 (0.9) | 15 (0.5) | 35 (0.4) |  |

The demographic and clinical characteristics of the study population at ART initiation were compared by three age-groups 10-14 years, 15-19 years, and 20-24 years. The three age-groups were similar in terms of pregnancy status and TB status at baseline (p=0.06, p=0.27 respectively) but differed in other characteristics. As expected, young adolescents (10-14years) were predominantly single (p<0.00), with the highest level of education at the primary level (p<0.00). Compared to the other age-groups, young adolescents also had higher enrollment at a tertiary healthcare facility (p<0.00), presented earlier for treatment (Median CD4 399cells/ML with mostly mild or no immune deficiency at baseline). Predominant ART regimen at baseline among young adolescents was ABC or AZT/3TC/EFV or NVP, compared to the other age-groups that were predominantly initiated on TDF/3TC/EFV or NVP. Older adolescents and young adult ART enrollees were almost exclusively female (82.9-88.1%), commonly married (42.9-59.8%), and educated up to a secondary or higher level of education (43.8-47.6%)

### Retention in care and viral outcomes

The overall proportion of ALHIV who had viral load testing was higher among young adolescents compared to older adolescents and young adults (59.1% vs 49.5% and 51.3%). Of ALHIV who had viral load testing, the proportion with VL less than 1000 copies/mL was lower among younger adolescents compared to older adolescents and young adults (63.5% vs 81.2% and 88.3%) (Figure 1)

**Figure 1.**
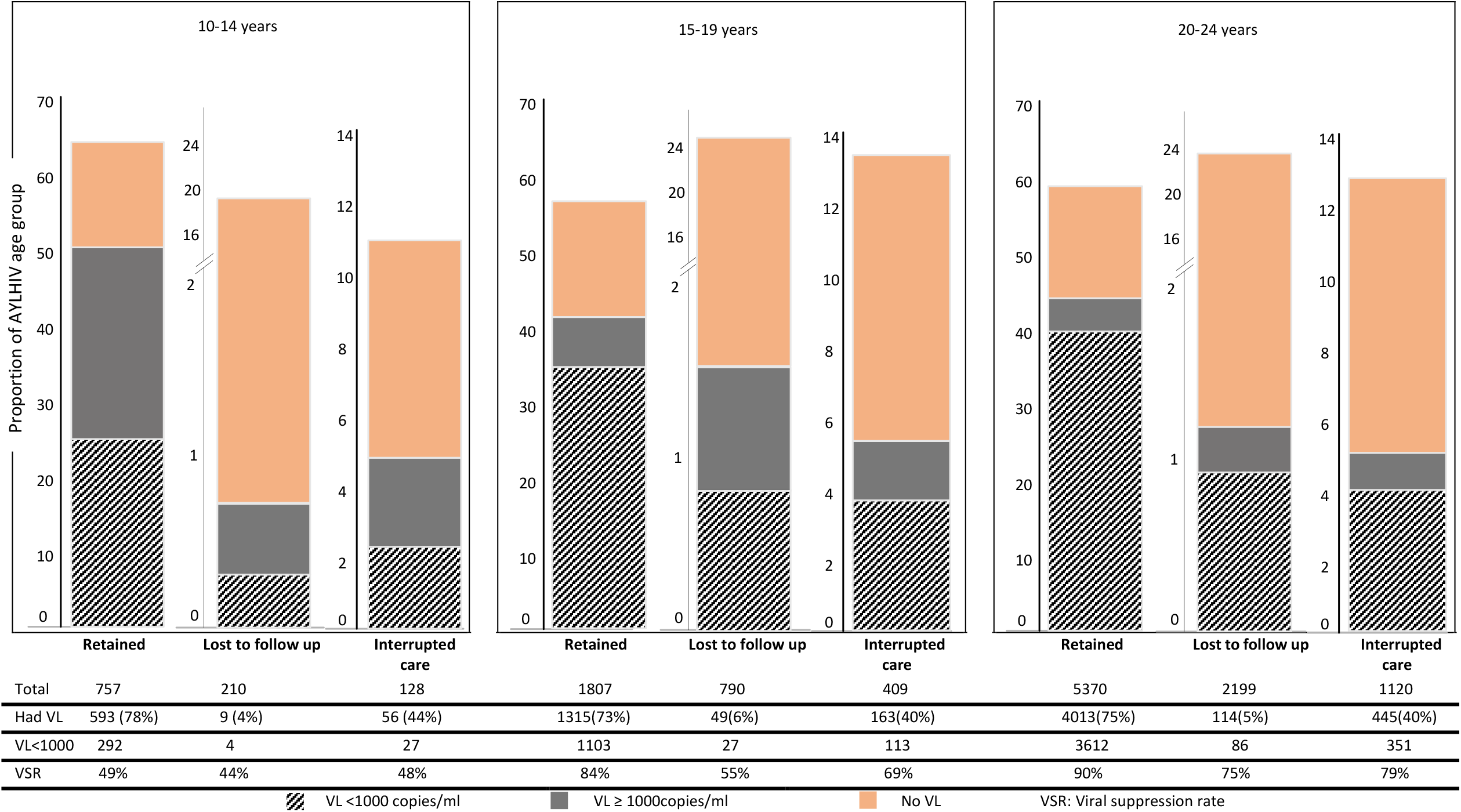
Retention in care and viral outcomes.

Viral load testing and viral suppression varied between age-groups by the pattern of care continuity. For ALHIV retained in care, the proportion who had viral load testing was highest among young adolescents (78%) compared to older adolescents (73%) and young adults (75%). Among retained ALHIV who had viral load testing, the viral suppression rate (the proportion that was virally suppressed) was lowest among young adolescents (49%) compared to older adolescents (84%) and young adults (90%).

For ALHIV lost to follow up, the proportion who had viral load testing was similar across the age-groups, ranging between 4-6%. Among ALHIV lost to follow up who had viral load testing young adolescents had the lowest viral suppression rate (44%) compared to older adolescents (55%) and young adults (75%).

For ALHIV who had interrupted care, the proportion who had viral load testing was highest among young adolescents (45%) compared to older adolescents (40%) and young adults (40%). Among ALHIV with interrupted care who had viral load testing young adolescents had the lowest proportion with viral suppression or viral suppression rate (48%) compared to older adolescents (69%) and young adults (79%).

### Measures of associations (fixed effects)

#### Continuity in care

Table 2 shows both fixed-effect (measures of association) and random-effects (measures of variation) from the multilevel analysis by age-group with non-retention in care as the outcome. Young adolescents: After controlling for both individual and facility characteristics the likelihood for non-retention was lower among young adolescents initiated on the ART regimen category ‘Others’ compared to young adolescents initiated on a regimen consisting of ABC or AZT/3TC/ ATVr or LPVr (aOR=0.25; p<0.05). The likelihood of non-retention in care reduced with increasing viral load testing coverage among young adolescents ALHIV. For each one-unit increase in health facility viral load test coverage, the odds of non-retention in this group decreased by 1% (aOR=0.99; p<0.001).

**Table 2:** Multilevel multivariable logistic regression of non-retention in care by age-group

| Characteristics | Young Adolescents(10-14years) |  |  | Older Adolescents (15-19years) |  |  | Young Adults (20-24 years) |  |  | Equality of Coefficient (p-value) |
| --- | --- | --- | --- | --- | --- | --- | --- | --- | --- | --- |
|  | Model 1 | Model 2 | Model 3 | Model 1 | Model 2 | Model 3 | Model 1 | Model 2 | Model 3 |  |
|  | OR | aOR | aOR | OR | aOR | aOR | OR | aOR | aOR |  |
| **LEVEL 1: INDIVIDUALS** |  |  |  |  |  |  |  |  |  |  |
| **Sex** |  |  |  |  |  |  |  |  |  | 5.02 (0.00) |
| Female |  |  |  |  |  |  |  |  |  |  |
| Male |  | 0.86 | 0.86 |  | 0.90 | 0.89 |  | 1.28** | 1.28** |  |
| **Marital Status** |  |  |  |  |  |  |  |  |  | 0.44(0.65) |
| Single |  |  |  |  |  |  |  |  |  |  |
| Married |  | 1.15 | 1.14 |  | 0.85 | 0.85 |  | 0.89 | 0.89 |  |
| Others |  | 1.13 | 1.12 |  | 0.91 | 0.90 |  | 0.97 | 0.96 |  |
| **Education level** |  |  |  |  |  |  |  |  |  | 0.11(0.89) |
| None/Informal |  |  |  |  |  |  |  |  |  |  |
| Primary |  | 0.89 | 0.89 |  | 1.03 | 1.03 |  | 1.01 | 1.00 |  |
| Secondary and higher |  | 1.01 | 1.01 |  | 0.92 | 0.92 |  | 0.95 | 0.94 |  |
| **Year of ART initiation** |  |  |  |  |  |  |  |  |  | 1.37(0.25) |
| 2015 |  |  |  |  |  |  |  |  |  |  |
| 2016 |  | 0.83 | 0.83 |  | 0.91 | 0.90 |  | 0.99 | 0.99 |  |
| 2017 |  | 0.85 | 0.86 |  | 0.75** | 0.74** |  | 0.91 | 0.91 |  |
| **WHO Stage** |  |  |  |  |  |  |  |  |  | 0.42(0.66) |
| 1 |  |  |  |  |  |  |  |  |  |  |
| 2 |  | 0.83 | 0.83 |  | 0.96 | 0.96 |  | 0.92 | 0.91 |  |
| 3 |  | 0.82 | 0.82 |  | 0.94 | 0.94 |  | 1.01 | 1.01 |  |
| 4 |  | 1.25 | 1.27 |  | 1.13 | 1.12 |  | 1.02 | 1.02 |  |
| **Baseline immunosuppression** |  |  |  |  |  |  |  |  |  | 0.23(0.79) |
| None (CD4≥500cells/ml) |  |  |  |  |  |  |  |  |  |  |
| Mild (CD4≥350-499cells/ml) |  | 1.09 | 1.09 |  | 1.03 | 1.03 |  | 1.08 | 1.08 |  |
| Advanced (CD4≥200-349cells/ml) |  | 1.24 | 1.24 |  | 1.02 | 1.03 |  | 1.09 | 1.09 |  |
| Severe (CD4 < 200cells/ml) |  | 1.31 | 1.31 |  | 1.13 | 1.15 |  | 1.17 | 1.17 |  |
| **Baseline TB status** |  |  |  |  |  |  |  |  |  | 0.11 (0.89) |
| No signs or symptoms of TB |  |  |  |  |  |  |  |  |  |  |
| Presumptive or confirmed TB |  | 1.01 | 1.01 |  | 0.96 | 0.94 |  | 1.02 | 1.01 |  |
| On IPT |  | 0.88 | 0.89 |  | 1.25 | 1.26 |  | 0.82 | 0.82 |  |
| **ART regimen at baseline** |  |  |  |  |  |  |  |  |  | 6.23 (0.00) |
| ABC or AZT/3TC/ ATVr or LPVr |  |  |  |  |  |  |  |  |  |  |
| ABC or AZT/3TC/EFV or NVP |  | 1.47 | 1.48 |  | 2.36 | 2.36 |  | 2.04 | 2.18 |  |
| TDF/3TC/EFV or NVP |  | 1.52 | 1.50 |  | 2.79 | 2.79 |  | 1.82 | 1.87 |  |
| TDF/3TC/ATVr or LPVr |  | 1.04 | 1.06 |  | 1.07 | 1.07 |  | 3.34 | 3.39 |  |
| Others |  | 0.23* | 0.25* |  | 1 | 1 |  | 0.14* | 0.15* |  |
| **LEVEL 2: Health Facility** |  |  |  |  |  |  |  |  |  |  |
| **Level of care** |  |  |  |  |  |  |  |  |  | 2.55(0.08) |
| Secondary |  |  |  |  |  |  |  |  |  |  |
| Tertiary |  |  | 1.10 |  |  | 0.93 |  |  | 1.38 |  |
| **Patient volume (AYLHIV)** |  |  |  |  |  |  |  |  |  | 1.48(0.23) |
| <200 |  |  |  |  |  |  |  |  |  |  |
| 200-400 |  |  | 1.07 |  |  | 1.20 |  |  | 1.29 |  |
| >500 |  |  | 1.03 |  |  | 0.65 |  |  | 0.67 |  |
| **Designated AYFS facility** |  |  |  |  |  |  |  |  |  | 1.38 (0.25) |
| Yes |  |  |  |  |  |  |  |  |  |  |
| No |  |  | 0.85 |  |  | 1.04 |  |  | 0.93 |  |
| **Viral load testing coverage** |  |  | 0.99** |  |  | 0.93** |  |  | 0.98** | 0.79(0.45) |
| **Random effects model** |  |  |  |  |  |  |  |  |  |  |
| Health Facility level |  |  |  |  |  |  |  |  |  |  |
| Variance estimate | 0.47** | 0.45** | 0.35** | 0.52** | 0.50** | 0.43** | 0.54** | 0.54** | 0.45** |  |
| Explained variation (%) | Ref | 4.26<sup>a</sup> | 22.22<sup>b</sup> | Ref | 3.84<sup>c</sup> | 14<sup>d</sup> | Ref | 0<sup>e</sup> | 16.67<sup>f</sup> |  |
| ICC (%) | 0.12 | 0.12 | 0.10 | 0.14 | 0.13 | 0.12 | 0.14 | 0.14 | 0.12 |  |

\* p<0.05; \*\* p<0.00
Model 1 is the null model; baseline model without any variable
Model 2 is model adjusted for individual-level variables,
Model 3 is additionally adjusted for facility-level covariates
a: Explained facility-level variance due to individual factors alone = (.47-.45)/.47 = 4.26%
b: Explained facility-level variance due to facility characteristics= (.45-.35)/.45 = 22.22%
c: Explained facility-level variance due to individual factors = (.52-.50)/.52 = 3.84%
d: Explained facility-level variance due to facility characteristics= (.50-.43)/.50 = 14%
e: Explained facility-level variance due to individual factors alone = (.54-.54)/.54 = 0%
f: Explained facility-level variance due to facility characteristics= (.54-.45)/.54 = 16.67%

Older adolescents: In this group non-retention was less likely among those enrolled in 2017 compared to enrollees in 2015 (aOR=0.74; p<0.001); no significant association between retention and any other individual-level factor was observed. At the level of health facility, the likelihood of non-retention in care reduced with increasing viral load testing coverage among enrolled AYLHIV. For each one-unit increase in health facility viral load test coverage, the odds of non-retention among older adolescents decreased by 7% (aOR=0.93; p<0.001).

Young adults: In this group, the odds of non-retention was higher among males (aOR=1.28; p<0.001) compared to females; the odds for non-retention was lower among those initiated on the ART regimen category ‘Others’ compared to those initiated on regimen ABC or AZT/3TC/ATVr or LPVr (aOR=0.15; p<0.05). At the level of the health facility, the likelihood of non-retention in care reduced with increasing viral load testing coverage among enrolled AYLHIV. For each one-unit increase in health facility viral load test coverage, the odds of non-retention among young adults decreased by 2% (aOR=0.98; p<0.001).

#### Viral suppression

Table 3 shows both fixed-effect (measures of association) and random-effects (measures of variation) from the multilevel analysis by age-group with viral suppression as the outcome.

**Table 3:** Multilevel multivariable logistic regression of viral suppression by age-group

| Characteristics | Young Adolescents(10-4years) |  |  | Older Adolescents (15-19years) |  |  | Young Adults(20-24 years) |  |  | Equality of Coefficient (p-value) |
| --- | --- | --- | --- | --- | --- | --- | --- | --- | --- | --- |
|  | Model 1 | Model 2 | Model 3 | Model 1 | Model 2 | Model 3 | Model 1 | Model 2 | Model 3 |  |
|  | OR | aOR | aOR | OR | aOR | aOR | OR | aOR | aOR |  |
| **LEVEL 1: INDIVIDUALS** |  |  |  |  |  |  |  |  |  |  |
| **Sex** |  |  |  |  |  |  |  |  |  | 1.12(0.33) |
| Female |  |  |  |  |  |  |  |  |  |  |
| Male |  | 0.73 | 0.73 |  | 0.69* | 0.69* |  | 0.93 | 0.94 |  |
| **Marital Status** |  |  |  |  |  |  |  |  |  | 0.67(0.51) |
| Single |  |  |  |  |  |  |  |  |  |  |
| Married |  | 1.77 | 1.77 |  | 1.74* | 1.7* |  | 1.36 | 1.35 |  |
| Others |  | 1.23 | 1.23 |  | 1.44 | 1.4 |  | 1.03 | 1.03 |  |
| **Education level** |  |  |  |  |  |  |  |  |  | 0.02(0.98) |
| None/Informal |  |  |  |  |  |  |  |  |  |  |
| Primary |  | 0.85 | 0.85 |  | 0.73 | 0.73 |  | 0.69 | 0.69 |  |
| Secondary and higher |  | 0.82 | 0.82 |  | 0.76 | 0.76 |  | 0.74 | 0.73 |  |
| **Year of ART initiation** |  |  |  |  |  |  |  |  |  | 0.7(0.49) |
| 2015 |  |  |  |  |  |  |  |  |  |  |
| 2016 |  | 1.54 | 1.54 |  | 0.81 | 0.81 |  | 1.13 | 1.15 |  |
| 2017 |  | 1.43 | 1.4 |  | 0.99 | 0.99 |  | 1.02 | 1.04 |  |
| **WHO Stage** |  |  |  |  |  |  |  |  |  | 0.21(0.81) |
| 1 |  |  |  |  |  |  |  |  |  |  |
| 2 |  | 0.79 | 0.79 |  | 0.79 | 0.80 |  | 1.04 | 1.04 |  |
| 3 |  | 1.73 | 1.73 |  | 0.69 | 0.70 |  | 0.78 | 0.78 |  |
| 4 |  | 0.60 | 0.60 |  | 0.82 | 0.80 |  | 0.87 | 0.90 |  |
| **Baseline immunosuppression** |  |  |  |  |  |  |  |  |  | 0.14(0.87) |
| None (CD4≥500cells/ml) |  |  |  |  |  |  |  |  |  |  |
| Mild (CD4≥350-499cells/ml) |  | 1.18 | 1.1 |  | 0.82 | 0.82 |  | 0.91 | 0.91 |  |
| Advanced(CD4≥200-349cells/ml) |  | 0.89 | 0.89 |  | 0.69 | 0.69 |  | 0.70 | 0.70 |  |
| Severe (CD4 < 200cells/ml) |  | 0.64 | 0.64 |  | 0.55* | 0.54 * |  | 0.55* | 0.55 ** |  |
| **Baseline TB status** |  |  |  |  |  |  |  |  |  | 0.22 (0.80) |
| No signs or symptoms of TB |  |  |  |  |  |  |  |  |  |  |
| Presumptive or confirmed TB |  | 0.69 | 0.69 |  | 1.19 | 1.18 |  | 1.160 | 1.19 |  |
| On IPT |  | 0.83 | 0.83 |  | 0.77 | 0.79 |  | 0.89 | 0.92 |  |
| **ART regimen at baseline** |  |  |  |  |  |  |  |  |  | 0.58(0.56) |
| ABC or AZT/3TC/ ATvr or LPVr |  |  |  |  |  |  |  |  |  |  |
| ABC or AZT/3TC/ EFV or NVP |  | 2.19 | 2.19 |  | 1.46 | 1.47 |  | 3.19 | 3.20 |  |
| TDF/3TC/EFV or NVP |  | 3.66 | 3.66 |  | 3.49 | 3.55 |  | 6.4* | 6.47 * |  |
| TDF/3TC/ATvr or LPVr |  | 0.15 | 0.15 |  | 2.25 | 2.27 |  | 0.74 | 0.81 |  |
| Others |  | 0.99 | 0.99 |  | 1.31 | 1.34 |  | 5.2 | 5.4 |  |
| **Continuity of care** |  |  |  |  |  |  |  |  |  | 0.1 (0.90) |
| Retained |  |  |  |  |  |  |  |  |  |  |
| LTF |  | 0.35 | 0.35 |  | 0.19 ** | 0.19** |  | 0.35 ** | 0.35** |  |
| Interrupted care |  | 0.40** | 0.42** |  | 0.41 ** | 0.41** |  | 0.42 ** | 0.42** |  |
| **LEVEL 2: Health facility** |  |  |  |  |  |  |  |  |  |  |
| **Level of care** |  |  |  |  |  |  |  |  |  | 0.39 (0.68) |
| Secondary |  |  |  |  |  |  |  |  |  |  |
| Tertiary |  |  | 0.90 |  |  | 0.90 |  |  | 0.80 |  |
| **Patient volume (AYLHIV)** |  |  |  |  |  |  |  |  |  | 0.12 (0.89) |
| <200 |  |  |  |  |  |  |  |  |  |  |
| 200-400 |  |  | 1.24 |  |  | 1.02 |  |  | 1.14 |  |
| >500 |  |  | 0.77 |  |  | 0.89 |  |  | 0.93 |  |
| **Designated AYFS facility** |  |  |  |  |  |  |  |  |  | 0.91(0.40) |
| No |  |  |  |  |  |  |  |  |  |  |
| Yes |  |  | 1.33 |  |  | 0.92 |  |  | 1.06 |  |
| **Viral load testing coverage** |  |  | 0.99 |  |  | 1.00 |  |  | 1.10 | 1.57(0.21) |
| **Random effects model** |  |  |  |  |  |  |  |  |  |  |
| Health facility level |  |  |  |  |  |  |  |  |  |  |
| Variance estimate (Facility level) | 0.39* | 0.38* | 0.32 | 0.49** | 0.44 ** | 0.44 ** | 0.34** | 0.29 ** | 0.28 ** |  |
| Explained variation (%) | Ref | 2.56<sup>a</sup> | 15.79<sup>b</sup> | Ref | 10.2<sup>c</sup> | 0<sup>d</sup> | Ref | 14.71<sup>e</sup> | 3.45<sup>f</sup> |  |
| Intra-facility ICC | 0.11 | 0.10 | 0.09 | 0.13 | 0.12 | 0.12 | 0.09 | 0.08 | 0.08 |  |

\*p<0.05, \*\*p<0.00; Model 1 is the null model; baseline model without any variable; Model 2 is model adjusted for
individual-level variables, Model 3 is additionally adjusted for facility-level covariates
a: Explained facility-level variance due to individual factors alone = (.39-.38)/.39 = 2.56%
b: Explained facility-level variance due to facility characteristics= (.38-.32)/.38 =15.79 %
c: Explained facility-level variance due to individual factors = (.49-.44)/.49 = 10.2%
d: Explained facility-level variance due to facility characteristics= (.44-.44)/.44 = 0%
e: Explained facility-level variance due to individual factors alone = (.34-.29)/.34 = 14.71%
f: Explained facility-level variance due to facility characteristics= (.29-.28)/.29 = 3.45%

Young adolescents: After controlling for both individual and facility-level characteristics, young adolescents who interrupted care were less likely to achieve viral suppression, compared to those retained (aOR=0.4; p<0.001). No facility-level characteristic was significantly associated with viral suppression in the age group.

Older adolescent ALHIV: The odds for viral suppression was higher among married, compared to single older adolescents (aOR=1.74; p<0.05). Compared to older adolescents with no immune suppression at baseline, those with severe immune suppression at baseline had a reduced likelihood of achieving viral suppression (aOR= 0.54; p<0.05). Compared to older adolescents retained in care, the odds for viral suppression was lower among older adolescents who were lost to follow up (aOR=0.19; p<0.001) or who interrupted care (aOR=0.41; p<0.001).

Young adults: Compared to young adults with no immune suppression at baseline, those with severe immune suppression at baseline had a reduced likelihood of achieving viral suppression (aOR= 0.55; p<0.001). Young adults initiated on ART regimen TDF/3TC/EFV or NVP had higher odds of achieving viral suppression compared to young adults initiated on ABC or AZT/3TC/ ATVr or LPVr (aOR= 6.47; p<0.05). Compared to young adults retained in care, the odds for viral suppression was lower among those who were lost to follow up (aOR=0.35; p<0.001) or who interrupted care (aOR=0.42; p<0.001).

### Measures of variations (random effects)

#### Retention in care

The results of the null models (Model 1) across ALHIV age-groups showed significant variance of 12-14% in the log-odds of non-retention in care attributed to health facilities [τ =0.47;p<0.0001 (young adolescents), τ =0.52; p<0.0001 (older adolescents), τ = 0.54; p<0.0001 (young adults)]. The variations at health facility level remained statistically significant even after controlling for individual-level (Model 2) and facility-level characteristics (Model 3), thereby lending support for the use of multilevel modeling to account for variations at the two levels. As judged by the proportional change in variance among young adolescents 4.26% of the variance in retention across health facilities was explained by individual factors (Model 2) and 22.223% of the variation explained by facility factors (Model 3). For older adolescents, 3.84% of this variance in retention was explained by individual factors alone (Model 2) and 14% explained by facility factors (Model 3). For young adults, none of the variations in retention was explained by individual factors alone (Model 2) while 16.67% was explained by both individual and facility factors (Model 3)

#### Viral suppression

The results of the null models (Model 1) across ALHIV age-groups show significant variance of 9-13% in the log-odds of viral suppression attributed to health facilities [τ =0.39; p<0.05 (young adolescents), τ =0.49; p<0.0001 (older adolescents), τ = 0.34; p<0.0001 (young adults)]. The variations at the health facility level also remained statistically significant even after controlling for individual-level and facility-level factors. As judged by the proportional change in variance among young adolescents 2.56% of the variance in viral suppression across health facilities was explained by individual factors alone (Model 2) and 15.79% of the variation explained by facility factors (Model 3). For older adolescents, 10.2% of this variance in viral suppression was explained by individual factors alone (Model 2), while facility factors did not explain any of the variance (Model 3). For young adults, 14.71% of the variation was explained by individual factors alone (Model 2) and 3.45% by facility factors (Model 3).

#### Multiple-group analysis

We detected statistically significant group differences in the association between retention in care and two variables: sex and ART regimen at baseline (Table 2).

The association between sex and retention in care differed in magnitude and direction of effects across the age-groups, reaching statistical significance only among young adults. with male young adults having an increased likelihood of not being retained in care (aOR 1.28, p<0.00).

The association between ART regimen at baseline and retention in care had similar direction but differed in the magnitude of effects across the age-groups. Initiation of regimen baseline ‘Others’ was associated with a reduced likelihood of not being retained in care in the three age-groups, with effect highest among young adults and lowest among older adolescents.

We did not find any significant group differences in the association between any of the explanatory variables and viral suppression (Table 3).

## 4. Discussion

In this multi-level and multiple group study, we estimated the relative contributions of individual-level and facility-level determinants to retention in care and viral suppression across ALHIV age-groups. We showed similarities and differences in magnitude and direction of effects across age-groups for individual level (age group, sex, ART regimen, and pattern of care continuity) and facility-level factors (viral load testing coverage) associated with retention in care and viral suppression. We discuss these findings according to the levels of influence.

### Individual-level factors

Although we found differences in the magnitude and direction of effects for the individual-level variables, two factors emerged as consistent predictors across age-groups: Baseline ART regimen and pattern of retention.

Similar to other study findings (8,11,12,18), higher proportions (64.7%) of young adolescents were retained in care compared to older adolescents (56.4%) and young adults living with HIV (58.7%). Despite their higher rates of retention, young adolescents had comparatively much lower viral suppression rates (63.5%) compared to older adolescents (81.2%) and young adults (88.3%). Also, while the odds for viral suppression reduced with lower levels of retention with increasing age, there was no significant difference in odds for viral suppression among young adolescents retained in care and those who were lost to follow-up. Although these findings seem counterintuitive since better retention in care is expected to lead to higher rates of viral suppression, it corroborates findings from other studies (30–35) showing high rates of viral non-suppression among young adolescents despite perfect adherence, most notably among young adolescents initiated on AZT + 3TC + NVP/EFV. Young adolescents in our study were initiated predominantly on ART regimen AZT + 3TC + NVP/EFV(63.2%), in contrast to older adolescents and young adults who were initiated predominantly on TDF/3TC/EFV or NVP (83.9 and 90.1% respectively). Moreover, the likelihood of achieving viral suppression irrespective of age-groups was highest with initiation on TDF/3TC/EFV or NVP regimen, reaching significance only among young adults ALHIV. These findings suggest a rise in regimen-specific disparity in viral outcomes, with increasing rates of viral non-suppression despite levels of adherence that should normally guarantee viral suppression. Such increases if not addressed could undermine other HIV prevention interventions.

Another reason we considered for the differential viral suppression between our age groups may be that substantial age-group differences exist by mode of HIV infection. Although we did not have enough information to characterize our cohort by mode of HIV infection, we speculate that the majority of younger adolescents in our study are adolescents perinatally infected with HIV compared to older adolescents and younger adults who we assume to be mostly behaviorally infected. Studies have shown that perinatally infected ALHIV are unique in their life-long experience with HIV infection, with many of them born in the era of mono- and dual-antiretroviral therapy, thus facing an increased likelihood of drug resistance (5,8,11,12)

We observed ART regimen specific effect differences on retention in care. Compared to ALHIV initiated on regimen ABC or AZT/3TC/ ATVr or LPVr, ALHIV initiated on other regimens than the previous ones, were less likely to be LTF or have interrupted care (i.e. not retained) in the three age groups. We speculate these differences to be due to drug side effects from ART which studies have shown to be a major barrier to adherence. Studies have suggested that, compared to patients who initiate ART after experiencing prior sicknesses, patients feeling healthy before commencing ART can have less motivation to tolerate the side effects associated with ART at different stages of care. The scenario is more likely to trigger disengagement with care among ALHIV who in the era of test and treat are generally more likely to initiate ART feeling healthy. The current HIV policy climate of “test and start” where laboratory and clinical assessments are no longer eligibility requirements for ART initiation may lead to an under-emphasis on clinical assessment processes including the management of side effects in favor of quicker ART initiations (36). While our study shows the benefit of early ART initiation to viral outcomes among ALHIV (i.e. likelihood of achieving viral suppression reduced with increasing immune-suppression at baseline, reaching statistical significance among older adolescent and young adults with severe baseline immune suppression) this finding points to an increased need to support ALHIV in balancing immediate needs and desires to live their normal lives with meeting the biomedical requirements for achieving viral suppression (clinic visits and treatment adherence).

We observed a preponderance of female ALHIV among our cohort with age, suggesting that female adolescents and young adult women represent a behaviorally vulnerable population. This supports previous findings from sub-Saharan Africa countries, where being young, female, and less-educated increases the chances for transgenerational HIV transmission (36). Combined with high adolescent fertility rates in Nigeria, poor retention, and low viral suppression rates among older adolescents and young adults in our study might undermine ongoing efforts at eliminating HIV mother-to-child transmission. However, we also found other gender-specific disparities within our cohort in which male young adult ALHIVs had a higher likelihood of interrupting care or being LTF, and male older adolescent ALHIV being less likely to achieve viral suppression.

Our findings on sex differences suggest the need to factor in gender considerations as services become increasingly differentiated for ALHIV. Studies have shown promising practices among the different approaches targeting men/male for improved HIV testing, prevention, treatment, care and support services(37–47)

### Health facility level factors

We found significant variation in outcomes attributable to differences at the health facility level. The variations across facilities remained statistically significant, even after controlling for individual- and facility-level factors (Models 2 and 3).

Although compared to individual-level factors facility-level characteristics accounted for a higher proportion of the total variance observed, the magnitude of this facility-level contribution to variance (i.e. facility effects) differed between ALHIV age-groups. For retention in care, higher increases due to facility effects were observed among young adolescents and young adults than among older adolescents. For viral suppression, there was no change in variance from facility effect, only a modest increase among young adults and a much higher increase in young adolescents. These differences suggest that interventions at the level of health services may be less effective than those targeting individual-level determinants for improving viral outcomes among older adolescents. This supports other studies showing that adolescent-focused health services interventions, though desirable, are not uniformly effective across all adolescent age-groups (48).

We found that an increase in viral-load testing at facility-level reduced the likelihood of non-retention among ALHIV. This might be explained by the high benefit perception of PLHIV concerning laboratory indicator measures (e.g. CD4, viral load test) for health status assessment. Some studies have shown that inability to access such tests often triggers disengagement with care (49–51). As the role of CD4 counts become increasingly outdated with the implementation of test and treat policies, increased coverage of routine viral load testing is necessary to encourage and sustain this positive health-seeking behavior. Furthermore, given disproportionately high rates of virological failure and sub-optimal retention among ALHIV, scale-up for viral load testing would allow providers to make informed decisions on counseling, support, and the management of treatment regimens(52).

### Clinical implications

Taken together, our study shows the need for both individual and health services interventions for ALHIV to be both broad-based and targeted. We showed that some factors at the individual and facility level (ART regimen, the pattern of retention in care, baseline immune suppression, viral load testing coverage) appear to be important predictors of outcomes in all ALHIV age-groups. These common factors appear to represent the base of a hierarchy of factors driving outcomes across all ALHIV and might be important entry points for modifying health services to better respond to ALHIV needs as part of a minimum package of services. This would include improving responsiveness to gender and age-specific needs, improving capacity for active case-finding and monitoring for retention in care, increased capacity for viral load testing, capacity building for all care providers to provide clinical care including pharmacovigilance, and monitoring of ART side effects.

### Study limitations

Our findings should be interpreted in light of some important limitations. Although we imputed for missing information to minimize bias, the possibility of non-differential errors cannot be ruled out. Due to differences in the age-group cohort sizes, there was greater power to detect covariate effect sizes among older adolescents and young adults age-groups, compared to young adolescents. Although case-finding was conducted for those ALHIV who were not retained in care, the percentage of these patients who returned to clinical care and had viral load assessment was low and may not be representative of their respective categories. We also lacked information to categorize our adolescent cohort by modes of transmission (perinatal vs behavioral infected) and previous exposure to antiretrovirals. Despite our use of multiple imputations to address missing data, we still cannot completely rule out some bias in our estimates given that health facilities with more missing data may also be the ones with poorer outcomes on the aggregate. Lastly, our study was carried out within the context of healthcare facilities receiving PEPFAR support. Findings may therefore not be generalizable to other ALHIV receiving care at health facilities not supported by PEPFAR who may differ in patient outcomes. Despite attempting to delineate the different levels of effects in our multilevel modeling our study still did not fully account for variations in outcomes at facility levels. This unobserved heterogeneity suggests the existence of other important explanatory factors that were not measured in our study.

### Conclusions

Our study supports our hypothesis that important differences exist in health service conditions between healthcare facilities that can explain variations in treatment outcomes among ALHIV. We also demonstrate key individual(ART regimen, baseline immune suppression, sex) and facility-level factors (facility-level viral-load testing) that may serve as entry points for both broad-based and targeted interventions to improve outcomes among ALHIV. Despite limited data to explore the whole range of factors at individual and service delivery levels potentially affecting treatment continuity and viral load suppression, we demonstrate a need to better understand and unpack these potentially modifiable factors at the individual- and facility-level to design interventions that ensure equity of among all ALHIV groups and ensure that no one is left behind.

## Data Availability

Data are not publicly available but can be obtained upon request from APIN-PEPFAR clinical database

## Authors’ contributions

OAB conceived the study, conducted the analysis, and wrote all drafts of the manuscript. All authors provided critical reviews of the draft. All authors read and approved of the final draft

## Acknowledgments

The authors acknowledge enabling support from APIN management during the conduct of this study. We also thank program managers and health workers who provide services to people living with HIV. Finally, we thank all adolescents and youths living with HIV whose consent to having their information used for research purposes has made this study possible

